# Longitudinal profiles of plasma gelsolin, cytokines and antibody expression predict COVID-19 severity and hospitalization outcomes

**DOI:** 10.1101/2022.06.01.22275882

**Authors:** Meshach Asare-Werehene, Michaeline McGuinty, Agatha Vranjkovic, Yannick Galipeau, Juthaporn Cowan, Bill Cameron, Curtis L. Cooper, Marc-André Langlois, Angela M. Crawley, Benjamin K. Tsang

**Affiliations:** Departments of Obstetrics & Gynecology, Immunity & Inflammation, University of Ottawa, Ottawa, Ontario, Canada, K1H 8L1; Departments of Cellular & Molecular Medicine, Immunity & Inflammation, University of Ottawa, Ottawa, Ontario, Canada, K1H 8L1; Departments of Biochemistry, Microbiology & Immunology, Immunity & Inflammation, University of Ottawa, Ottawa, Ontario, Canada, K1H 8L1; Departments of Medicine, Immunity & Inflammation, University of Ottawa, Ottawa, Ontario, Canada, K1H 8L1; Departments of Centre for Infection, Immunity & Inflammation, University of Ottawa, Ottawa, Ontario, Canada, K1H 8L1; Chronic Disease Program, Ottawa Hospital Research Institute, Ottawa, Canada, Ottawa, Ontario, Canada K1H 8L6; Clinical Epidemiology Program, Ottawa Hospital Research Institute, Ottawa, Canada, Ottawa, Ontario, Canada K1H 8L6; Department of Biology, Carleton University, Ottawa, Ontario, Canada; Coronavirus Variants Rapid Response Network–Biobank, Canada

## Abstract

**Background:** Prognostic markers for COVID-19 disease outcome are currently lacking. Plasma gelsolin (pGSN) is an actin-binding protein and an innate immune marker involved in disease pathogenesis and viral infections. Here, we demonstrate the utility of pGSN as a prognostic marker for COVID-19 disease outcome; a test performance that is significantly improved when combined with cytokines and antibodies compared to other conventional markers such as CRP and ferritin.

**Methods:** Blood samples were longitudinally collected from hospitalized COVID-19 patients as well as COVID-19 negative controls and the levels of pGSN in μg/mL, cytokines and anti-SARS-CoV-2 spike protein antibodies assayed. Mean±SEM values were correlated with clinical parameters to develop a prognostic platform.

**Results:** pGSN levels were significantly reduced in COVID-19 patients compared to healthy individuals. Additionally, pGSN levels combined with plasma IL-6, IP-10 and M-CSF significantly distinguished COVID-19 patients from healthy individuals. While pGSN and anti-spike IgG titers together strongly predict COVID-19 severity and death, the combination of pGSN and IL-6 was a significant predictor of milder disease and favorable outcomes.

**Conclusion:** Taken together, these findings suggest that multi-parameter analysis of pGSN, cytokines and antibodies could predict COVID-19 hospitalization outcomes with greater certainty compared with conventional clinical laboratory markers such as CRP and ferritin. This research will inform and improve clinical management and health system interventions in response to SARS-CoV-2 infection.

**Trial Registration:** N/A

**Funding:** The Ottawa Hospital Department of Medicine - Special Pandemic Agile Research Competition

## Introduction

COVID-19 is a respiratory infectious disease caused by SARS-CoV-2 (1-4). Currently, >460 million people are infected worldwide and >6 million SARS-CoV-2-related deaths have been recorded (World Health Organization) thus, requiring urgent health care interventions. SARS-CoV-2 suppresses the adaptive arm (B and T lymphocytes) of the immune system and overly activates the innate arm (macrophages, natural killer cells, dendritic cells), causing an inflammatory cytokine storm (TNF-α, IL-6, IL-8, IL-1β, MCP-1) (5). These immune reactions can result in lung damage, multiple organ dysfunctions and in severe cases, death (5, 6). To date, direct therapeutic options are limited to a modestly effective antiviral that remains inaccessible to most patients. Patients are managed using best supportive care including steroids, and in severe cases the use of mechanical ventilation and extracorporeal membrane oxygenation (ECMO) (7, 8). Identifying novel biomarkers will enable physicians to predict/assess the progress of COVID-19 patients and inform decision-making for available therapeutic intervention.

Lower viral titers contribute to false negative testing for SARS-CoV-2 (9, 10). Thus, an additional screening method that could detect possible infections despite lower viral titers will be highly beneficial. Antibodies directed to the nucleoprotein of SARS-COV-2 could be useful in identifying recent infections given most of the population will have detectable anti-spike antibody from prior infection and/or immunization (9, 10). As to whether their detection could be used to determine COVID-19 disease severity and clinical outcomes remain to be examined.

Laboratory markers including interleukin 6 (IL-6), C-reactive protein (CRP), ferritin, platelets, lactate dehydrogenase (LDH), erythrocyte sedimentation rate (ESR), D-dimer and lymphopenia are significantly implicated in COVID-19 complications (11, 12). Diseases like influenza, bacterial infection, and chronic inflammatory and neurodegenerative diseases trigger the elevation of these laboratory markers (13-15), hence casting doubts on their specificity and sensitivity as reliable prognostic markers for COVID-19 severity, hospitalization outcomes and ICU admissions.

Recently, proteomic screening of plasma samples from individuals with active COVID-19 disease has shown that plasma gelsolin (pGSN) expression levels are downregulated compared to healthy controls (16-18). Gelsolin (GSN) is a 80-85kDa calcium-dependent multifunctional actin-binding protein which exists in three isoforms: cytoplasmic gelsolin (cGSN), pGSN, and gelsolin 3 (19, 20). pGSN has recently been shown to be a key player in the immune regulation of cancer (21). pGSN depletion significantly reduces its organ-protective function, leading to multi-organ dysfunction syndrome (MODS), increased mortality and long-term morbidity in survivors, complications that are commonly seen in COVID-19 patients (22-25).

Thus, it is conceivable that pGSN could be of clinical relevance in the pathogenesis of COVID-19. Its pathological significance and potential as a biomarker for COVID-19 severity and treatment response remains to be determined. In the present study, we have demonstrated a pGSN multi-analyte panel as a potential biomarker of poor COVID-19 outcomes and pGSN with cytokines and antibodies present as a significant predictor of COVID-19 hospitalization outcomes compared with conventional clinical laboratory markers.

## Results

### pGSN’s combination with IL-6, IP-10 and M-CSF significantly distinguish hospitalized COVID-19 patients from healthy controls

COVID-19 negative and positive patients were recruited at TOH and their blood longitudinally collected. The subjects were stratified into COVID-19 negative and positive subjects and the levels of pGSN, cytokines and antibodies were quantified **(Fig. 1A)**. There were no significant differences in the levels of pGSN between males and females as well as age references **(Suppl. Fig S1A and SB)**. pGSN levels were significantly lower (p<0.0001) in COVID-19 patients compared with the negative subjects **(Fig. 1B)**. On the contrary, IL-6 (p=0.034), M-CSF (p=0.001) and IP-10 (p=0.023) levels were significantly up-regulated in COVID-19 patients compared with the negative subjects whereas HGF and CTAK demonstrated a tendency to be significantly higher **(Fig. 1B)**. The ratio of pGSN and cytokines were calculated and used in several ratio analyses. The mean levels of pGSN/IL-6, pGSN/M-CSF, pGSN/HGF, pGSN/IP-10 and pGSN/CTAK were determined and compared between COVID-19 patients and COVID-19 negative controls. All ratios tested were significantly decreased in COVID-19 patients compared with COVID-19 negative controls **(Fig. 1C; p < 0.0001)**. ROC curves were used to determine the diagnostic significance of these ratios in differentiating COVID-19+ patients from COVID-19-controls **(Fig. 1D)**. The combination of pGSN with IL-6, IP-10 and M-CSF but not HGF provided a significant 100% test accuracy with AUC of 1 **(Fig. 1D)**. Meanwhile, pGSN alone or cytokines alone provided a statistically significant AUC <1 **(Fig. 1D)**. To determine if these markers predicted disease progression in COVID-19 patients, pGSN levels measured over time were compared between days of sample collection. No significant differences were observed over time between the groups; however, a significant improvement **(p<0.002)** in the levels of pGSN were observed on day 30, potentially suggesting patients’ conditions might be improving **(Fig. 1E’ and Suppl. Figs. S1C-G)**.

**Figure 1.**
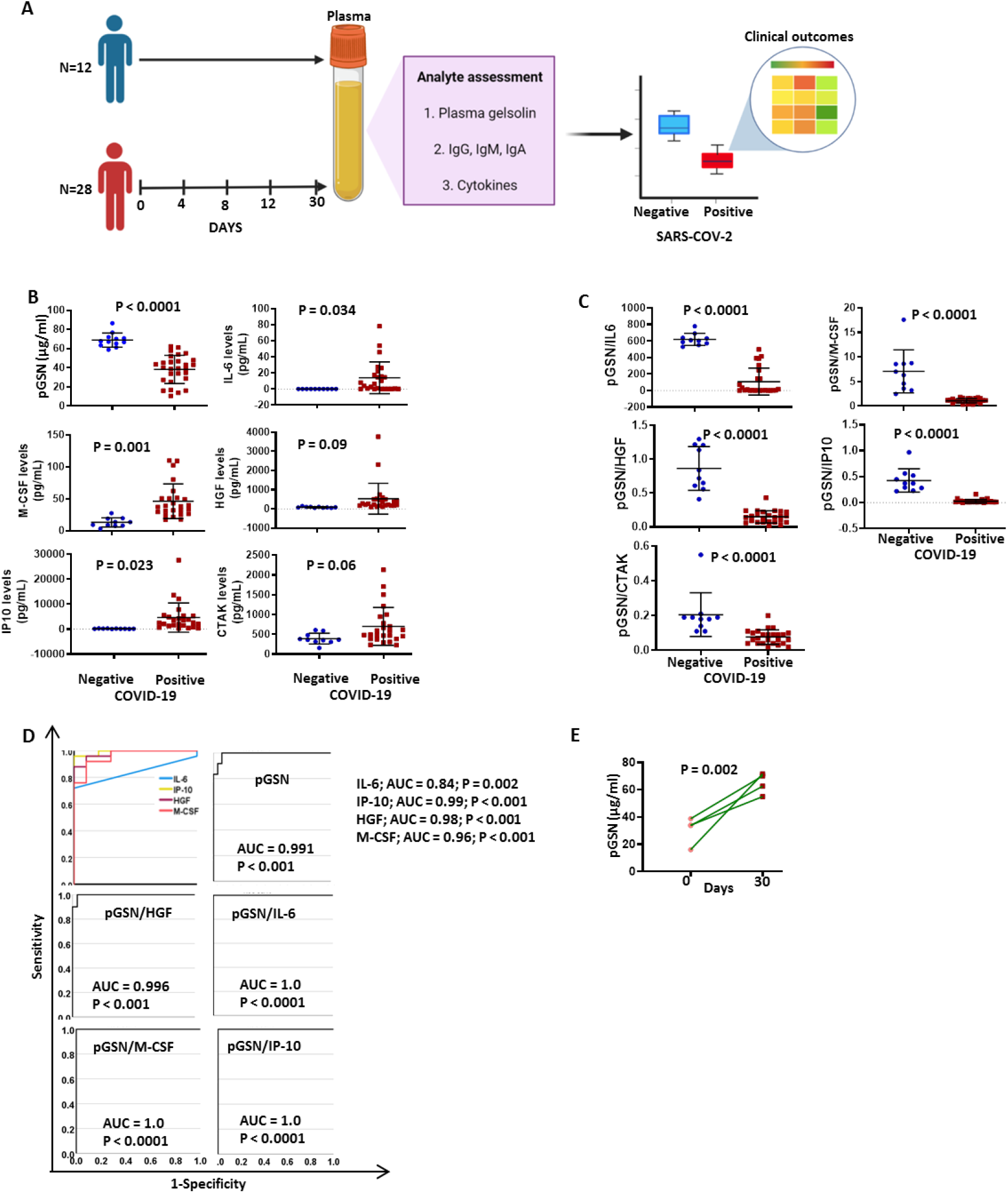
pGSN’s combination with IL-6, IP-10 and M-CSF significantly discriminates COVID-19 positive patients from negative subjects. (A) 28 samples were serially collected from PCR-confirmed SARS-CoV-2-infected patients and 12 samples collected from non-COVID-19 controls. Samples were analyzed for the levels of pGSN, cytokines and anti-SARS-COV-2 antibodies. The mean levels of the analyte were compared between COVID-19 negative and positive subjects. The test performances of the biomarkers against clinical outcomes were analyzed. (B) The mean±SEM levels of pGSN, IL-6, IP-10, M-CSF, HGF and CTAK were detected and compared between COVID-19 and non-COVID-19 patients. (C) The ratio of pGSN and cytokines were calculated and used as multi-analyte diagnostic panels. The mean±SEM levels of pGSN/IL-6, pGSN/M-CSF, pGSN/HGF, pGSN/IP-10 and pGSN/CTAK were determined and compared between COVID-19 and non-COVID-19 subjects. (D) The test performances of pGSN, cytokines (IL-6, IP-10, HGF, M-CSF) and multi-analyte panels (pGSN/HGF, pGSN/IL-6, pGSN/M-CSF and pGSN/IP-10) in differentiating COVID-19 patients from non-COVID-19 subjects were determined using receiving operating characteristic (ROC) curves. (E) pGSN levels in blood collected on day 0 and 30 were compared. P-values were calculated by independent sample t-test.

### The combination of pGSN and IL-6, IP-10, HGF, CTAK and M-CSF is associated with favorable COVID-19 outcomes

Having observed that pGSN significantly distinguishes COVID-19**+** patients from COVID-19 negative controls, its prediction of disease severity, progression and hospitalization outcomes were subsequently investigated. Here, we correlated the levels of pGSN at day 1 of hospitalization with the clinical outcomes of the patients prior to being discharged alive or death **(Fig. 2)**. We observed that pGSN levels increased with increases in ICU admissions **(Fig. 2A)**. COVID-19 patients who were discharged had increased levels of pGSN suggesting their recovery **(Fig. 2A)**. Inflammatory cytokines (IL-6, IP-10, HGF, M-CSF and CTAK) as well as pGSN were elevated in severe COVID-19 patients reminiscent of reported cytokine storms (6, 26, 27) that are highly associated with increased organ damage **(Fig. 2B)**.

**Figure 2.**
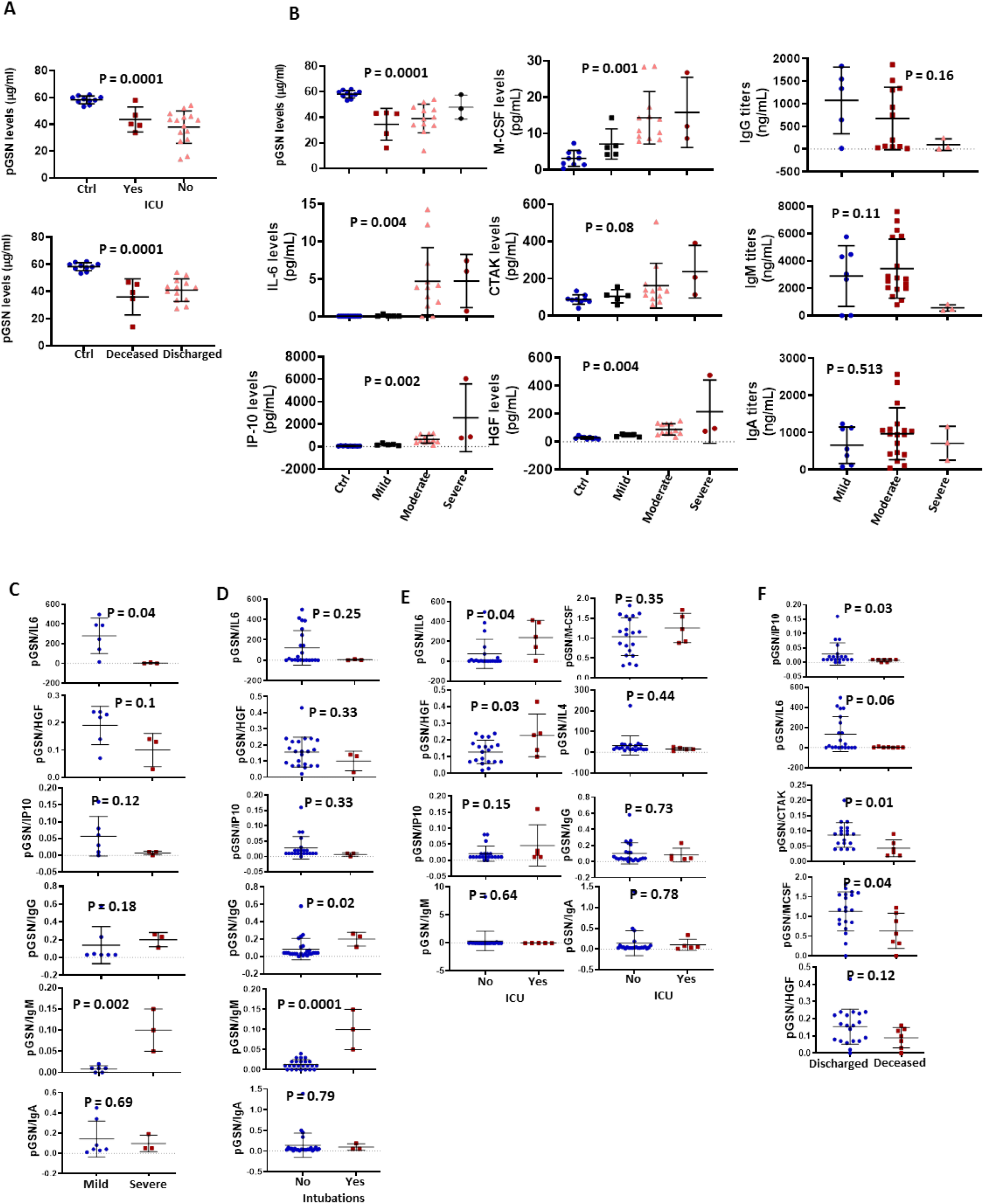
The combination of pGSN and IL-6, IP-10, HGF, CTAK and M-CSF is associated with favorable COVID-19 outcomes. (A) pGSN levels at day 0 were determined and the mean±SEM compared between patients based on ICU admittance and hospitalization outcome. (B) The mean±SEM levels of pGSN, IL-6, IP-10, M-CSF, HGF, CTAK, IgA, IgM and IgG were detected and compared between patients based on disease severity. The ratio of pGSN and cytokines were calculated and used as multi-analyte diagnostic panels. (C) The mean±SEM levels of pGSN/IL-6, pGSN/HGF, pGSN/IP-10, pGSN/IgG, pGSN/IgM and pGSN/IgA were determined and compared between mild and severe COVID-19 patients as well as (D) intubation status and (E) ICU admittance with pGSN/IL-4 included. (F) The mean±SEM levels of pGSN/IP-10, pGSN/IL-6, pGSN/CTAK, pGSN/M-CSF and pGSN/HGF were compared between deceased and discharged patients. P-values were calculated by one-way ANOVA and independent sample t-test.

Unlike cytokines, none of the antibody isotypes measured associated with disease severity **(Fig. 2B)**. In order to develop a stronger prognostic marker, pGSN was combined with selected cytokines in a ratio to be compared between mild and severe cases, ICU admission and hospitalization outcomes. pGSN/IL-6 was significantly higher in mild cases compared with severe cases; pGSN/HGF and pGSN/IP-10 showed a similar trend although not significant **(Fig. 2C)**. Meanwhile, pGSN/IgM was significantly higher in severe cases with pGSN/IgG (but not pGSN/IgA) showing a similar trend **(Fig. 2C)**. Although the ratio of pGSN and cytokines and IgA had no significant association with intubation, pGSN/IgM and pGSN/IgG at day 1 were significantly elevated in patients who were intubated during hospitalization **(Fig. 2D)**. In terms of ICU admissions, patients with significantly higher levels of pGSN/IL-6 and pGSN/HGF (but not pGSN/IP-10, pGSN/IL-4 and pGSN/M-CSF) were admitted to the ICU **(Fig 2E)** with the antibodies having no significant impact. The levels of pGSN/IP-10, pGSN/CTAK, pGSN/IL-6 and pGSN/M-CSF (but not pGSN/HGF) were elevated in patients that were discharged **(Fig. 2F)**, suggesting that pGSN levels in combination with inflammatory cytokines especially pGSN/IL-6, may predict mild and favorable outcomes for hospitalized COVID-19 patients.

### The pGSN/IL-6 ratio is a strong predictor of disease progression and outcomes relative to symptoms onset

To further strengthen the potential of using pGSN and cytokines as a multi-analyte marker for COVID-19 outcomes, we investigated their utility in days from symptoms onset. Mild COVID-19 patients had significantly high pGSN/IL-6 amongst all the multi-analyte markers following day 4 of symptoms onset compared with moderate and severe COVID-19 patients, and pGSN/IP-10 and pGSN/HGF showed a trend toward this association **(Fig. 3A; Suppl. Fig. S2A)**. Patients admitted into the ICU as well as those who died had lower pGSN/IL-6, with pGSN/HGF showing a similar trend **(Fig. 2A; Suppl. Fig S2B-C)**. Taken together, the pGSN/IL-6 ratio presents as a strong candidate which could be potentially used to monitor disease progression and hospitalization outcomes 4 days from symptom onset. Although pGSN levels correlated with SARS-COV-2 spike protein antibodies on day 8, their combination as a multi-analyte marker did not provide a significant output for monitoring patient outcomes days from symptom onset **(Fig. 3B; Suppl. Figs. S2-S3)**.

**Figure 3.**
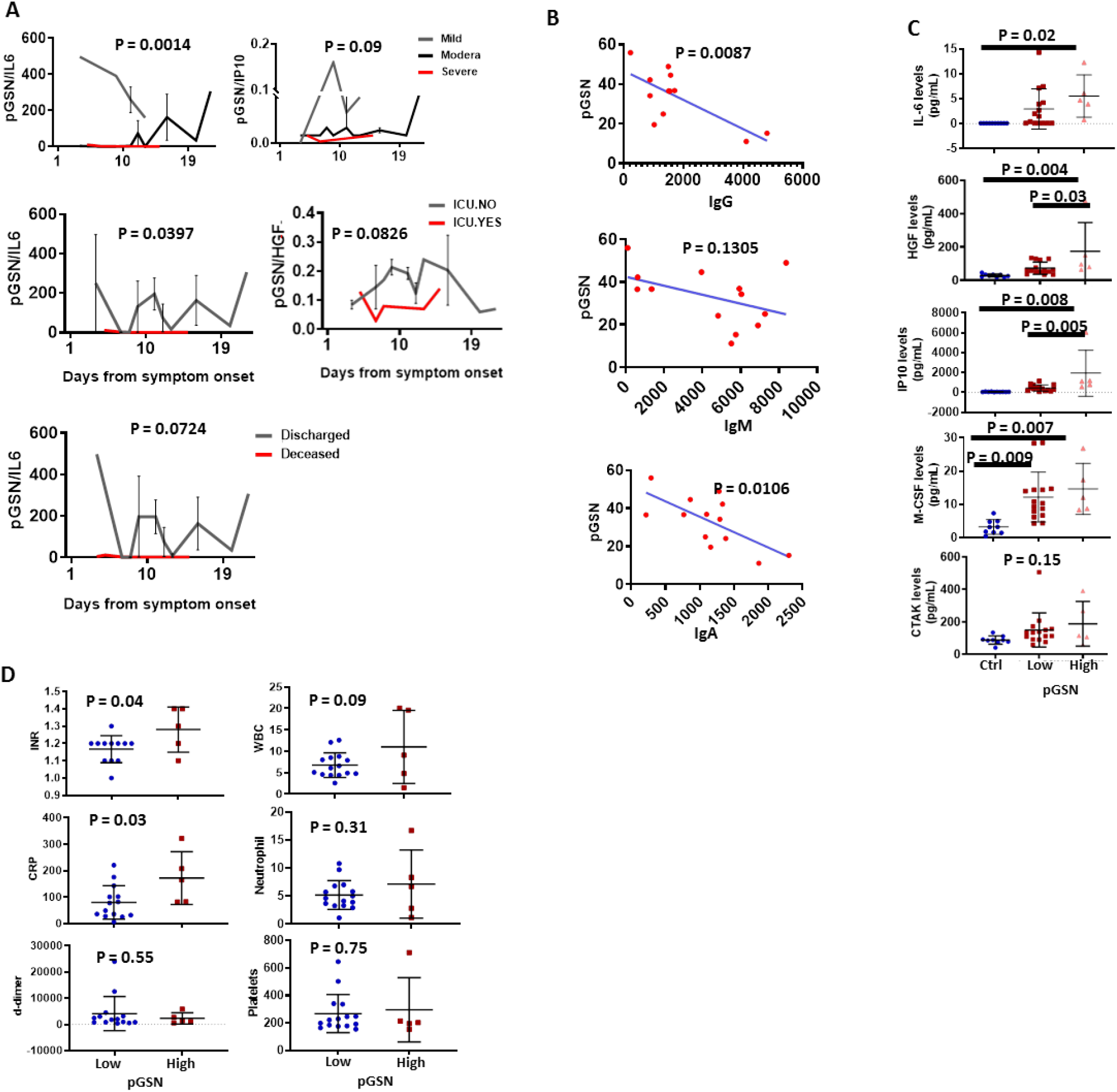
pGSN/IL-6 multi-analyte is a strong marker for monitoring disease severity and outcomes days from symptoms onset. (A) pGSN/IL-6, pGSN/IP-10 and pGSN/HGF levels were utilized to monitor disease severity, ICU admittance and discharge following day 1 from symptoms onset. (B) pGSN and SARS-COV-2 antibodies (IgG, IgM and IgA) were assessed on day 8 and correlated using Pearson’s test. (C) Patients were stratified into low and high pGSN groups using an optimal cut-off point. The mean±SEM levels of IL-6, HGF, IP-10, M-CSF and CTAK were compared between the groups using scatter plot. (D) The mean±SEM levels of INR, CRP, d-dimer, WBC, neutrophils and platelets were compared between the pGSN groups using scatter plot. P-values were calculated by one-way ANOVA and independent sample t-test.

### pGSN levels are elevated in response to cytokine storm

We have thus far demonstrated that pGSN and inflammatory cytokines are elevated in severe COVID-19 disease although we have yet to determine how they correlate with each other as well as that of the clinically used inflammatory markers. Here, we stratified patients into two groups based on their pGSN levels (low and high) using an optimal cut-off point determined by Fisher’s Exact Test. The mean inflammatory cytokine levels (IL-6, IP-10, HGF, M-CSF and CTAK; **Fig. 3C**) and clinically used laboratory markers **(INR, d-dimer, CRP, WBC, platelets, neutrophils; Fig. 3D)** were determined and compared between the groups. The mean levels of the cytokines (except CTAK) were significantly higher in the high pGSN group compared with lower pGSN group **(Fig. 3C)** whereas only INR and CRP were elevated in the high pGSN group. These findings suggest that pGSN could be upregulated as a feedback mechanism to fight the damage caused by the inflammatory state of the patient.

### pGSN/IgG and pGSN/IgM are strong predictors of worse COVID-19 outcomes

We further tested how reliable pGSN would be to monitor disease severity and hospitalization outcomes either alone or in combination with SARS-COV-2 spike antibodies at the time of hospitalization (day 0). Using ROC curves, we derived AUC to assess the prognostic performances of these markers **(Fig. 4)**. Using pGSN, CRP, IgG, IgM or IgA alone were not significantly predictive, however the AUCs of pGSN/IgG and pGSN/IgM to predict severe COVID-19 were 0.893 (p=0.028) and 0.96 (p=0.01) respectively **(Fig. 4A; Suppl. S4A)**. Similar outcomes were observed for pGSN/IgG (AUC=0.776; p=0.032) and pGSN/IgM (AUC=0.735; p=0.067) when predicting COVID-19 death **(Fig. 4B; Suppl. S4B)**. These findings suggest that pGSN/IgG and pGSN/IgM could be potential markers for predicting disease severity and hospitalization outcomes for COVID-19.

**Figure 4.**
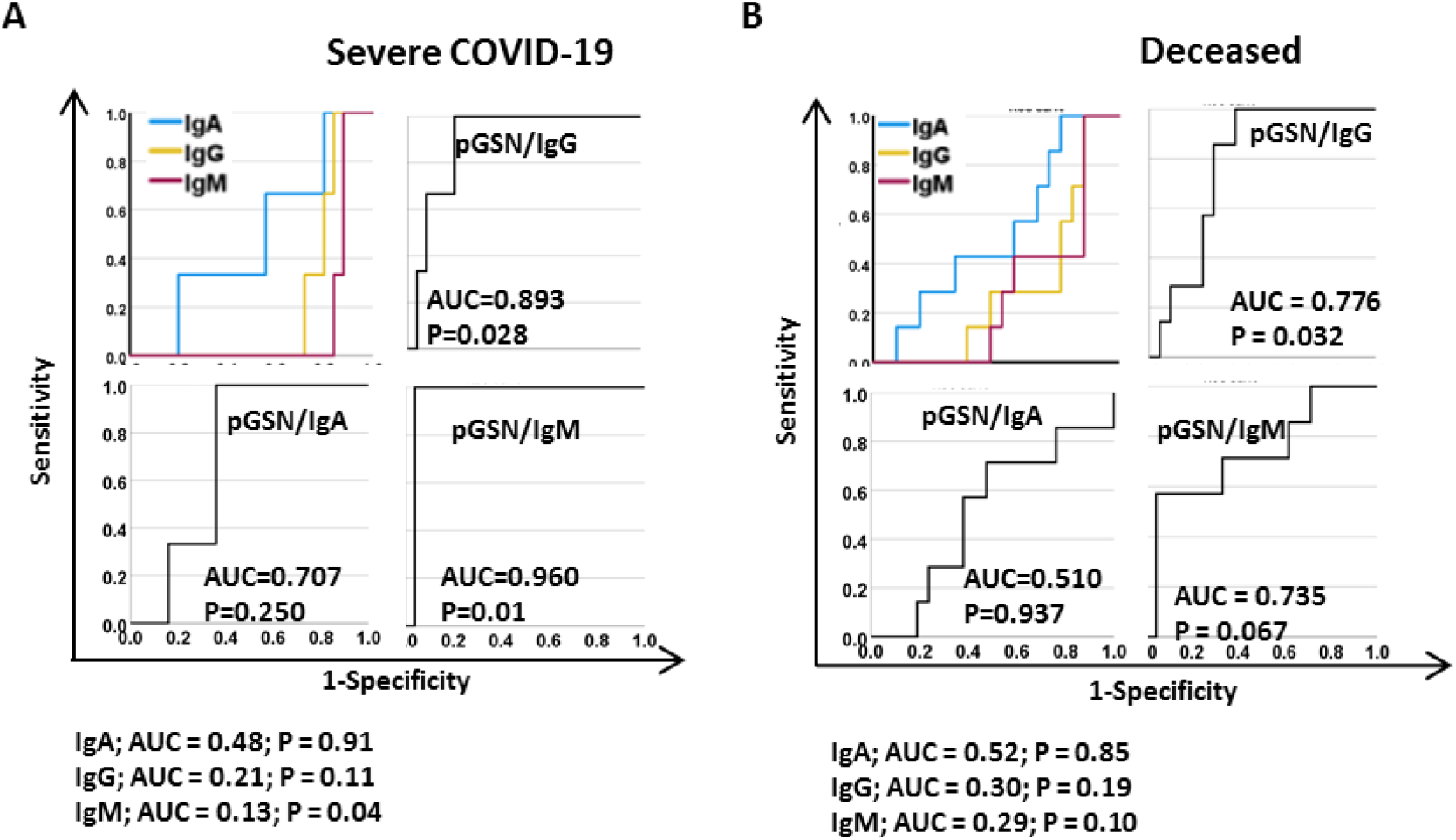
pGSN/IgG and pGSN/IgM are strong predictors of worst COVID-19 outcomes. ROC curves were used to assess the test performances of IgG, IgA, IgM, pGSN/IgG, pGSN/IgM and pGSN/IgA in predicting (A) disease severity and (B) death.

### pGSN/IL-6 is a strong predictor of mild COVID-19 and favorable outcomes

Unlike the antibodies, the inflammatory cytokines showed more promise when combined with pGSN for predicting mild and favorable COVID-19 outcomes (i.e. no-ICU admissions, and survival to hospital discharge). pGSN, CRP and individual cytokines alone provided an AUC < 0.6 in predicting mild COVID-19 disease **(Fig. 5A; Suppl. Fig. S1C)**. Meanwhile, combining pGSN with IL-6 (AUC=0.904; p=0.003) showed a more significant ability to predict mild COVID-19 and IP-10 (AUC=0.754; p=0.065), HGF (AUC=0.746; p=0.075) and TGF-β1 (AUC=0.765; p=0.078) demonstrated a trend toward significance **(Fig. 5A; Suppl. Fig. S4C)**.

**Figure 5.**
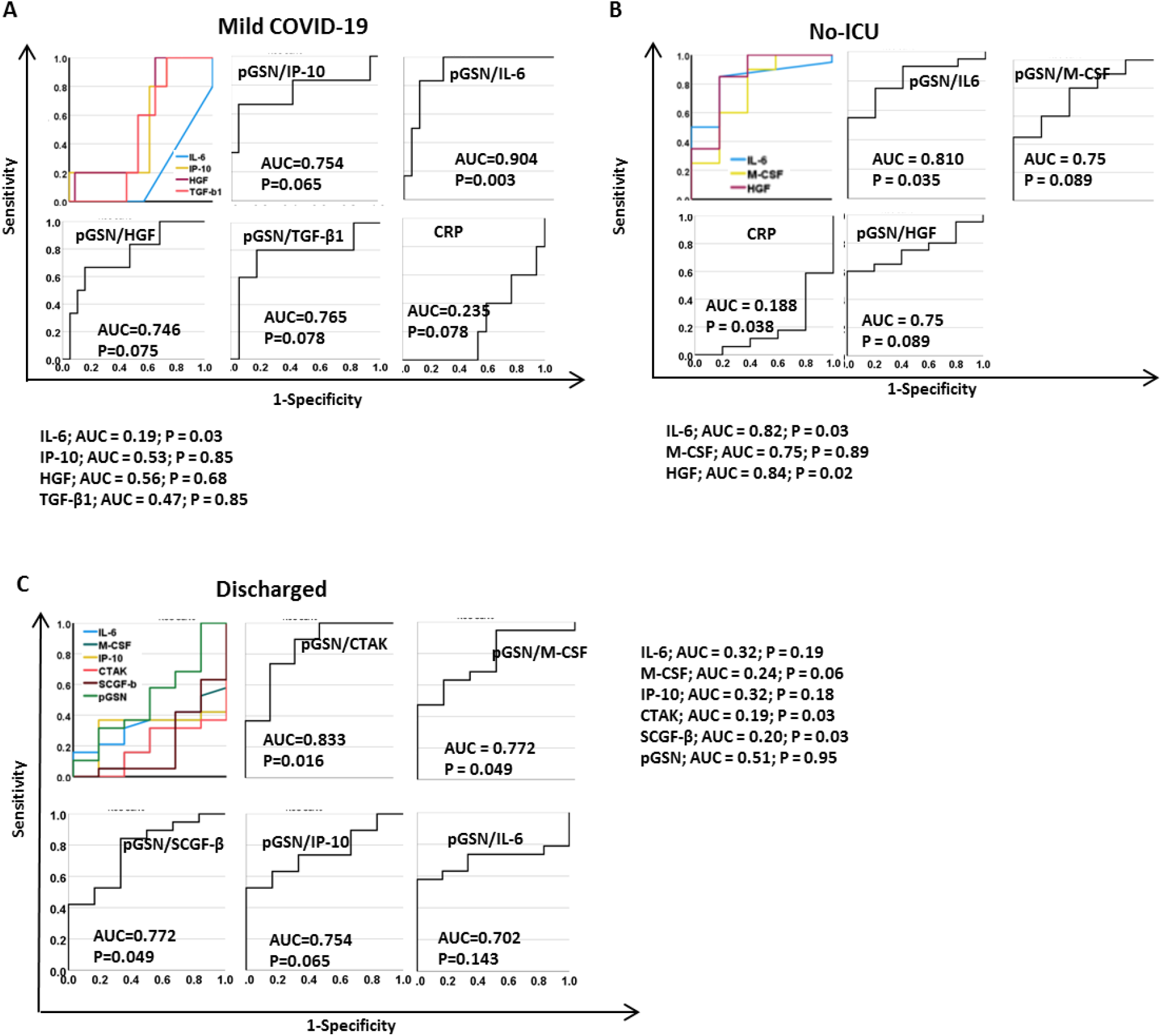
pGSN/IL-6 is a strong predictor of mild COVID-19 cases and favorable outcomes. ROC curves were used to assess the test performances of IL-6, IP-10, HGF, TGF-β1, M-CSF, CTAK, SCGF-β, CRP, pGSN, pGSN/IP-10, pGSN/IL-6, pGSN/HGF, pGSN/TGF-β, pGSN/M-CSF and pGSN/SCGF-β in predicting (A) mild COVID-19 cases, (B) no-ICU admissions and (C) discharged patients.

Unlike the mild cases, there were no significant differences between the pGSN/cytokine panels and pGSN alone when predicting non-ICU admissions **(Fig. 5B; Suppl. Fig. S4D)**. Interestingly, pGSN/CTAK (AUC=0.833; p=0.016), pGSN/M-CSF (AUC=0.772; p=0.049) and pGSN/SCGF-b (AUC=0.772; p=0.049) but not the other ratios, significantly predicted survival to hospital discharge of COVID-19 patients **(Fig. 5C)**. As expected, a modest AUC (less than 6) was observed with pGSN, cytokines or clinical inflammatory markers alone, suggesting the need for a multi-analyte marker for disease monitoring in COVID-19 **(Fig. 5C; Suppl. Fig. S4E)**.

### Multi-analyte panel of pGSN with cytokines and antibodies predict more than one clinical outcome of COVID-19

Using heat maps, we demonstrated how our multi-analyte panels could enable us to predict two clinical outcomes at the same time; an approach that will be significantly useful in triaging patients. We observed that COVID-19 patients with increased levels of pGSN/IL-6, pGSN/HGF and pGSN/IgM compared with the other markers are more likely not to progress to severe disease and will eventually be discharged alive **(Fig. 6A; Suppl. Fig. S5A)**. The situation with pGSN/IgG was different since it is more predictive of severe disease and death in this study **(Fig. 6A)**. Additionally, patients with increased levels of pGSN/IL-6, pGSN/IP-10 and pGSN/IgM compared with the other markers were more likely not to be admitted to the ICU and to survive to hospital discharge **(Fig. 6B; Suppl. Fig. S5B)**. When ICU and disease severity were used in the heat map analysis, we observed that patients with increased pGSN/IL-6, pGSN/IP-10 and pGSN/IgM are likely to present with mild cased and will eventually avoid ICU admission. These findings suggest that by combining pGSN with cytokines more than one clinical outcome could be predicted in COVID-19 cases from the first day of hospital visit.

**Figure 6.**
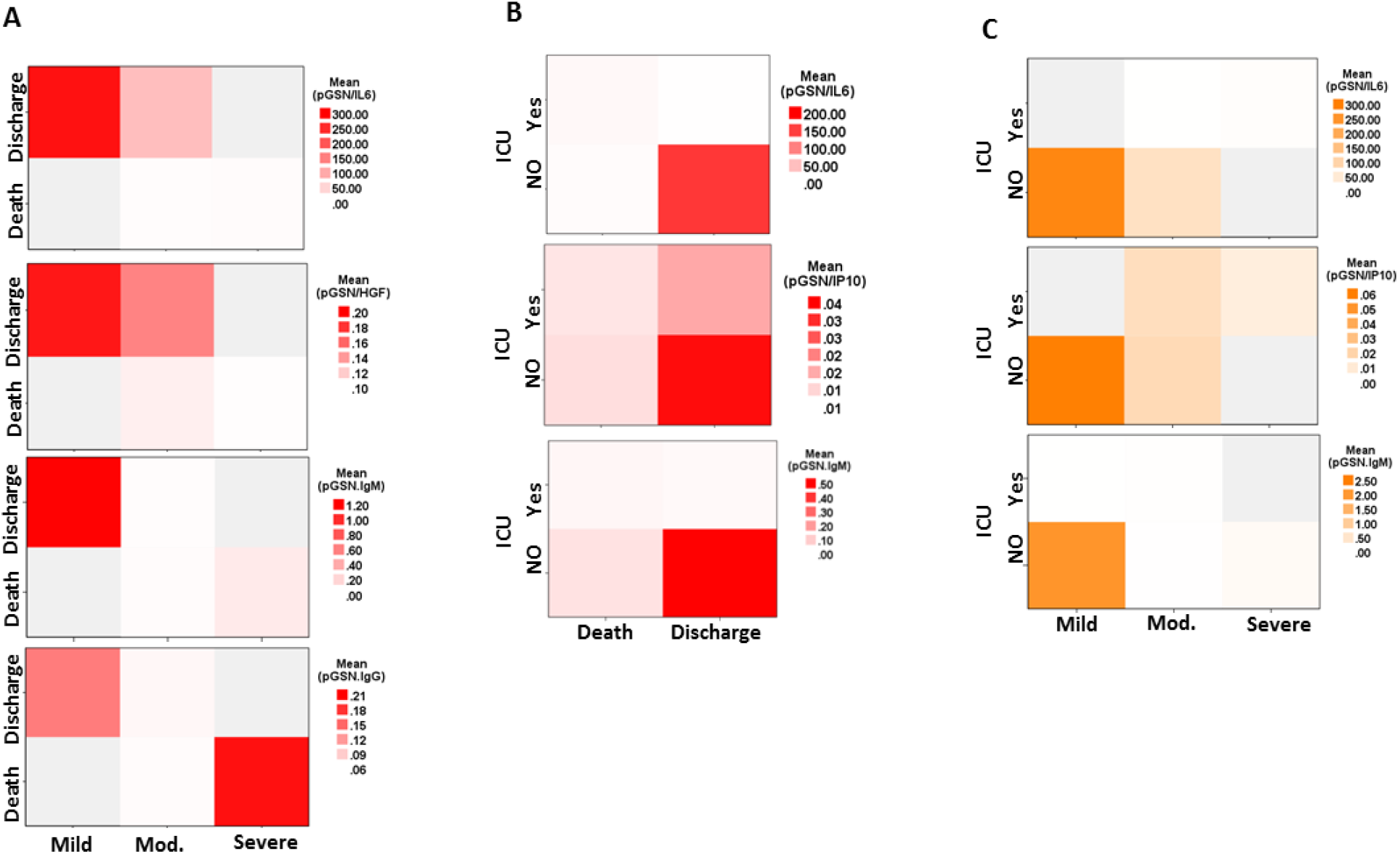
Multi-analyte panel of pGSN with cytokines and antibodies predict more than one clinical outcome of COVID-19. pGSN/IL-6, pGSN/HGF, pGSN/IP-10, pGSN/IgM and pGSN/IgG were used in a heat map analyses to predict (A) disease severity and hospitalization outcomes, (B) ICU admissions and hospitalization outcomes and (C) disease severity and ICU admissions.

## Discussion

Our study reveals for the first time the prognostic significance of pGSN when combined with cytokines and anti-SARS-CoV-2 antibodies in COVID-19 infection. Specifically, we have demonstrated that pGSN is significantly downregulated in COVID-19 patients and in combination with cytokines and anti-SARS-COV-2 antibodies can predict COVID-19 severity and hospitalization outcomes.

Although pGSN levels were lower at the time of admission, a return to normal range in the levels were seen at day 30 which is associated with improved outcome (i.e. discharge) in patients with COVID-19 infection. pGSN alone outperformed cytokines (IL-6, IP-10, HGF and M-CSF) in discriminating COVID-19+ patients from non-COVID-19 suggesting that pGSN is a potential biomarker indicative of COVID-19 infection. As shown in previous studies (5, 6, 28, 29), COVID-19 is characterized by the upregulation of pro-inflammatory cytokines which was evident in our study with the upregulation of IL-6, IP-10, M-CSF, HGF, CTAK and TGF-b1. It is interesting to note that the test accuracy of pGSN was enhanced when combined with IL-6, IP-10, HGF and C-MSF in a multi-analyte panel. During cytokine storm (5), multi-organ damage results in the release of actin filaments in the blood which are cleared, in part, by pGSN (30, 31), a process that may result in the decrease of pGSN in COVID-19 patients also. This might further explain why combining pGSN with the pro-inflammatory cytokines provided much greater accuracy since these markers are both systemically related to disease progression.

Prediction of hospitalization outcomes is key to the effective management of COVID-19 cases as well as allocation of hospital resources. However, there are currently no biomarkers which reliably predict COVID-19 severity, ICU admission and death. We have shown that a multi-analyte panel using pGSN and cytokines can predict favorable hospitalization outcomes. pGSN/IL-6 was the strongest predictive marker (amongst other cytokines) for mild COVID-19+ cases, ICU admissions and patient discharge. pGSN is a negative regulator of IL-6 and other pro-inflammatory cytokines secreted by monocytes and macrophages (32). Hence, pGSN could be secreted as a response to cytokine storm to prevent multiple organ damage as well as actin filament toxicity. This notion is consistent with the observation that pro-inflammatory cytokine levels were elevated in COVID-19+ patients with high pGSN. It is therefore note-worthy that combining pGSN and IL-6 provided robust test accuracy for favorable outcomes compared to individual markers alone, an indication of disease progression in the participants. Additionally, our findings are consistent with a previous study that has demonstrated the immunomodulatory effectiveness of human recombinant (rhu)-pGSN in the treatment of a single severe COVID-19 case (33). There is an ongoing double-blinded proof-of-concept trial (NCT04358406) to further assess the efficacy and safety of rhu-pGSN in severe COVID-19.

Another key observation in this study was the relationship between pGSN levels and antibodies (IgA, IgM and IgG) against SARS-CoV-2 spike protein. From day 1 of sample collection through day 30 we observed a pattern where the inverse relationship between pGSN and the antibodies increased. An indication that with improving clinical status, pGSN levels are restored while antibody levels, especially IgG, began to decline. This is consistent with the observation that antibody levels were lower in patients with severe COVID-19. Some patients were treated with corticosteroids; which could impact antibody levels. Unlike the pro-inflammatory cytokines, combining pGSN with the anti-spike IgG and IgM were strong predictive markers for severe COVID-19 and COVID-19-related death. Actin remodeling is a crucial step in B-cell activation and B-cell receptor function (34, 35). Although pGSN is a key protein in actin binding and remodeling (20, 30), we have yet to investigate whether it is directly involved in COVID-19 humoral immunity. This could explain the relationship between pGSN and the antibodies as observed in these COVID-19 patients. As to whether pGSN has a direct function on the production of SARS-COV-2 antibodies is yet to be investigated.

An additional major finding was the ability to utilize pGSN/IL-6 as well as other markers (cytokines, antibodies and clinical laboratory inflammatory markers) to predict more than one clinical outcome. Of note, we observed that COVID-19 patients with high pGSN/IL-6 and pGSN/IgM presented with mild disease did not require ICU admission and were more likely to be discharged. The opposite was noted with pGSN/IgG. In situations where there are limited resources, this could potentially provide clinicians and hospital administrators’ prior knowledge in prioritizing their resources such as ICU beds in terms of delivering care to COVID-19 patients. In summary, a multi-analyte panel of pGSN, cytokines and SARS-COV-2 antibodies, present as potential biomarkers predicting disease severity, ICU admissions and COVID-19-related death. Despite these promising findings, there are limitations that need to be acknowledged. The sample size of this study is relatively small due to the difficulty in sampling patients longitudinally in a hospital setting. Antibody kinetics (levels and isotype) during the early infection could be influenced by time since infection. Also, the control samples consist of out-patient populations that had no recorded history of respiratory infection during sample collection. Having control samples from patients who are SARS-CoV-2 negative but positive for other respiratory diseases will further strengthen the reliability of pGSN multi-analyte panels. In future studies, patient sample size will be increased while we investigate the prognostic significance of pGSN multi-analyte panel in SARS-CoV-2 variant patients, vaccinated patients as well as convalescent plasma.

## Materials and Methods

### Ethics statement

This study was approved by the Ottawa Health Science Network Research Ethics Board (OH SNREB; Protocol# 20200200-01H) and conducted in accordance with the appropriate guidelines. Written informed consent was obtained from all subjects.

### Patient characteristics and plasma samples

Forty participants (unvaccinated) recruited at The Ottawa Hospital (TOH) from 2020 to 2021 were included in this study. Plasma samples were serially collected from 28 PCR-confirmed SARS-CoV-2-infected patients admitted to hospital at days 0, 4, 8, 12 and 30. Plasma samples had pre-determined laboratory markers such as C-reactive protein (CRP), ferritin, white blood cells (WBC), d-dimer, neutrophils, platelets and international normalized ratio (INR). Twelve samples collected from healthy controls were negative for SARS-COV-2. Non-COVID-19 subjects had no reported illnesses at the time of sample collection. COVID-19 severity was defined as mild (no supplemental oxygen), moderate [supplemental oxygen, not high flow or no flow listed, no intubation/mechanical ventilation – ex. No Bi-level Positive Airway Pressure (BiPap) /intubation)] and severe (high flow oxygen and non-invasive or invasive mechanical ventilation – ex. Yes BiPap/intubation). The range and median of follow-up period was 87 and 10 days respectively. Other details of patient demographics and baseline characteristics are described in **Supplementary Table 1**. Patients received interventional management including antiviral therapies (lopinavir/ritonavir, ribavirin, hydroxychloroquine and remdesivir) and non-antiviral adjunctive therapies (convalescent plasma, steroids, immune-modulators, anticoagulation, and antibiotics) in addition to appropriate supportive care.

### pGSN ELISA

pGSN concentrations were assayed in the study participants by sandwich ELISA (Aviscera Bioscience, Inc. CA), according to manufacturer’s instructions. The detection antibody was raised against human plasma (soluble) gelsolin. Plasma samples were diluted in a sample buffer (1/1500; Aviscera Bioscience, Inc. CA) and all analyses done in triplicate. Optical densities (OD) were determined using a microtiter plate reader at 450 nm. The blank was subtracted from the triplicate readings for each standard and sample and concentrations reported in µg/mL.

### Cytokine Assay

The concentrations of pro- and anti-inflammatory cytokines in plasma were quantified using multiplexing immunobead assays analyzed using the BioRad Luminex machine (Bio-Rad Laboratories, Hercules, CA, USA). Transforming growth factor β (TFG-β) -1, -2 and -3 were quantified using the TGF-Beta 1,2,3 Magnetic Bead Kit (Milliplex, Millipore SIGMA) while all remaining cytokines were assayed using the 48-plex Bio-Plex Pro™ Human Cytokine Screening Panel (BioRad, Mississauga, ON, CANADA). Cytokine concentrations were reported in pg/mL.

### Anti-spike protein antibody assay

A manual colorimetric ELISA was used to measure antibodies targeting SARS-CoV-2 spike protein, a complete description of the methods can be found here (36). Briefly, high protein-binding Immulon 4 HBX clear 96-well plates (VWR International, Mississauga, ON, Canada, #62402-959) were coated with 50 μL of 2 μg/mL full length trimeric Spike protein (NRC) diluted in PBS and kept at 4°C on a shaker. After an overnight incubation, the plates were washed three times with 200uL of PBST and blocked with 200 μL of 3% w/v skim milk powder in PBST for 1 hour at room temperature on a shaker. During that time, plasma samples were diluted in 1% w/v skim milk powder. The plates were washed again and 100uL of diluted plasma samples was added to the plate. Additionally, positive and negative serum controls alongside an isotype-antigen specific calibration curve (CR3022 Human IgG1 (Absolute Antibody, Ab01680-10.0), anti-SARS-CoV-2 S CR3022 Human IgA (Absolute Antibody, Ab01680-16.0), or anti-SARS-CoV-2 S CR3022 Human IgM (Absolute Antibody, Ab01680-15.0)) was added to the plate and incubated for 2h with shaking. The plates were washed again and 50uL of isotype specific secondary antibodies (anti-human IgG#5-HRP (NRC), anti-human IgA-HRP (Jackson ImmunoResearch Labs, 109-035-011), and anti-human IgM-HRP (Jackson ImmunoResearch Labs, 109-035-129) added and incubated with shaking for 1 hour. The plates were then washed for one last time and developed using 100 μL of SIGMAFAST OPD Tablets (Sigma-Aldrich, P9187) dissolved in 20 mL of Water for Injection for Cell Culture (Thermo Fisher Scientific, A1287301). The reaction was stopped with the addition of 50uL of 3M HCl and the absorbance was measured at 490 nm using a PowerWave XS2 Plate Reader (BioTek Instruments). Absorbance reads were blank adjusted and correlated with the linear portion of the calibration curve, hereby taking into account plate-to-plate variability and dilution factor. Concentrations of antibodies were reported in ng/mL.

### Biostatistical methods

The GraphPad Prism 8 (San Diego, CA, USA) and SPSS software version 28 (SPSS Inc., Chicago, IL, USA) were used to perform all statistical analyses and two-sided P ≤ 0.05 considered to indicate statistical significance. Concentrations of pGSN in study groups were dichotomized by their median cut-offs into low or high concentrations and correlated to clinicopathological parameters (37) [age, sex, disease severity, inflammatory cytokines, IgG, IgA and IgM levels], using Pearson’s correlation test (two-tailed), Fisher exact test, Students’ *t* test and Kruskal Wallis test, as appropriate. Receiving operating characteristic (ROC) curves were used to assess the performance of pGSN, cytokines and multi-analyte panels over their entire range of values. The area under the curve (AUC) was used as an index of global test performance to predict disease severity. Heat maps were used to demonstrate the relationships between multi-analyte panels and clinical outcomes.

## Data Availability

All data produced in the present study are available upon reasonable request to the authors.

## Author Contributions

M.A-W. and B.K.T. conceived and designed the study in partnership with A.M.C. Plasma samples were collected by B.C., C.L.C. and M.M and processed by A.V., and A.M.C. Clinical information was collected and curated by B.C., C.L.C., and M.M. pGSN analyses were performed by M.A-W and B.K.T. Cytokine assays were conducted by A.V. and A.M.C. while antibody measurements were completed by Y.G. and MA.L. M.A.W. analysed the data with scientific input from B.K.T., B.C., C.L.C., MA.L., M.M., J.C. and A.M.C. M.A.W. wrote the paper with feedback from all authors.

## Acknowledgements

This work was supported by a grant from The Ottawa Hospital Department of Medicine - Special Pandemic Agile Research Competition (M.M., C.L.C., J.C., A.M.C.).

