## Supplemental Figures and Table for "Longitudinal profiles of plasma gelsolin, cytokines and antibody expression predict COVID-19 severity and hospitalization outcomes"

### Supp. Fig. S1

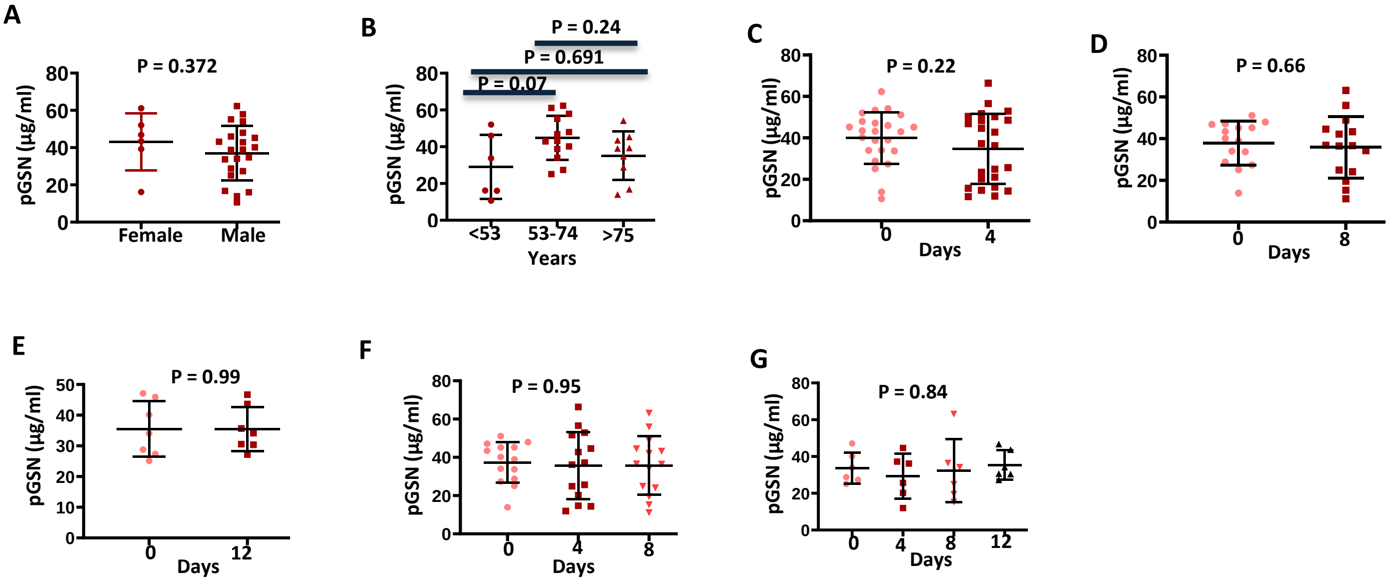


**Supp. Fig. S1. pGSN levels are not affected by gender and age.** The mean±SEM levels of pGSN were compared between **(A)** male and female COVID-19 patients as well as **(B)** age ranges. **(C-G)** The mean±SEM levels of pGSN were also compared between days of sample collection. *P*-values were calculated using one-way ANOVA and independent sample *t*-test.

### Supp. Fig. S2

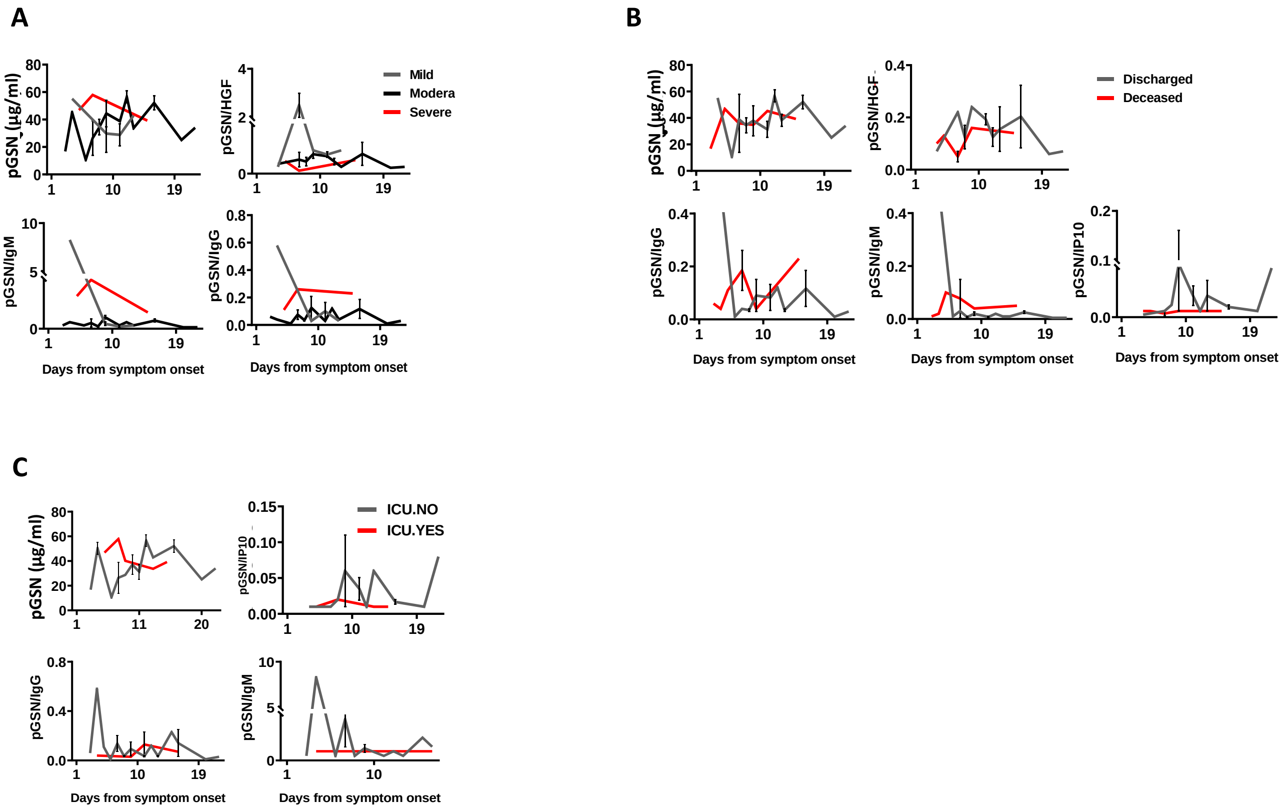


**Supp. Fig. S2. pGSN multi-analyte panels for monitoring disease severity and outcomes days from symptoms onset.** pGSN, pGSN/HGF, pGSN/IP-10, pGSN/IgM and pGSN/IgG were utilized to monitor **(A)** disease severity, **(B)** discharge and **(C)** ICU admittance following day 1 from symptoms onset. *P*-values were calculated by one-way ANOVA and independent sample *t*-test.

### Supp. Fig. S3

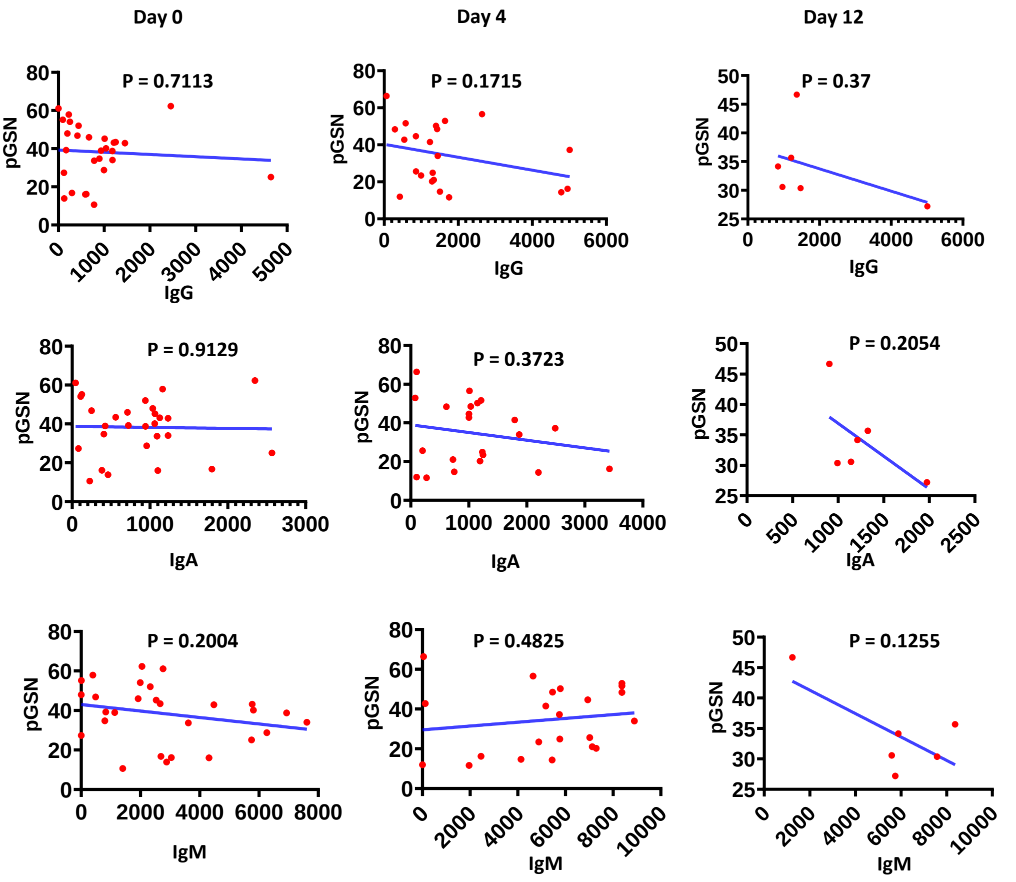


**Supp. Fig. S3. Correlation between pGSN and SARS-COV-2 antibodies.** pGSN and SARS-COV-2 antibodies (IgG, IgM and IgA) were assessed on day 0, 4 and 12 and correlated using Pearson's test.

### Supp. Fig. S4

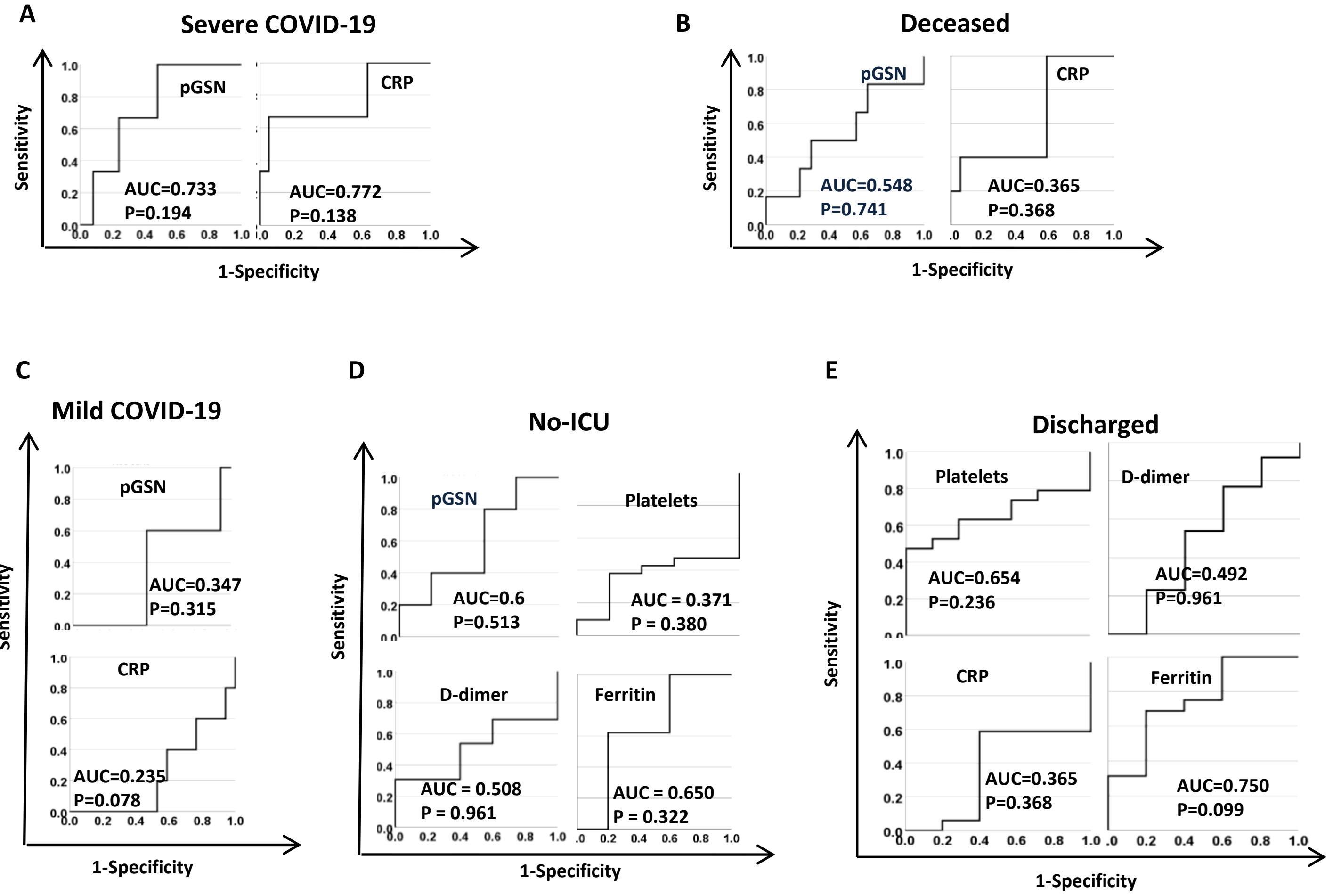


**Supp. Fig. S4. pGSN multi-analyte panels as predictors of COVID-19 outcomes.** ROC curves were used to assess the test performances of pGSN, CRP, platelets, D-dimer and Ferritin in predicting **(A)** severe COVID-19, **(B)** deceased patients, **(C)** mild COVID-19 cases, **(D)** ICU admissions and **(E)** patients that are discharged.

**Supp. Fig. 5**

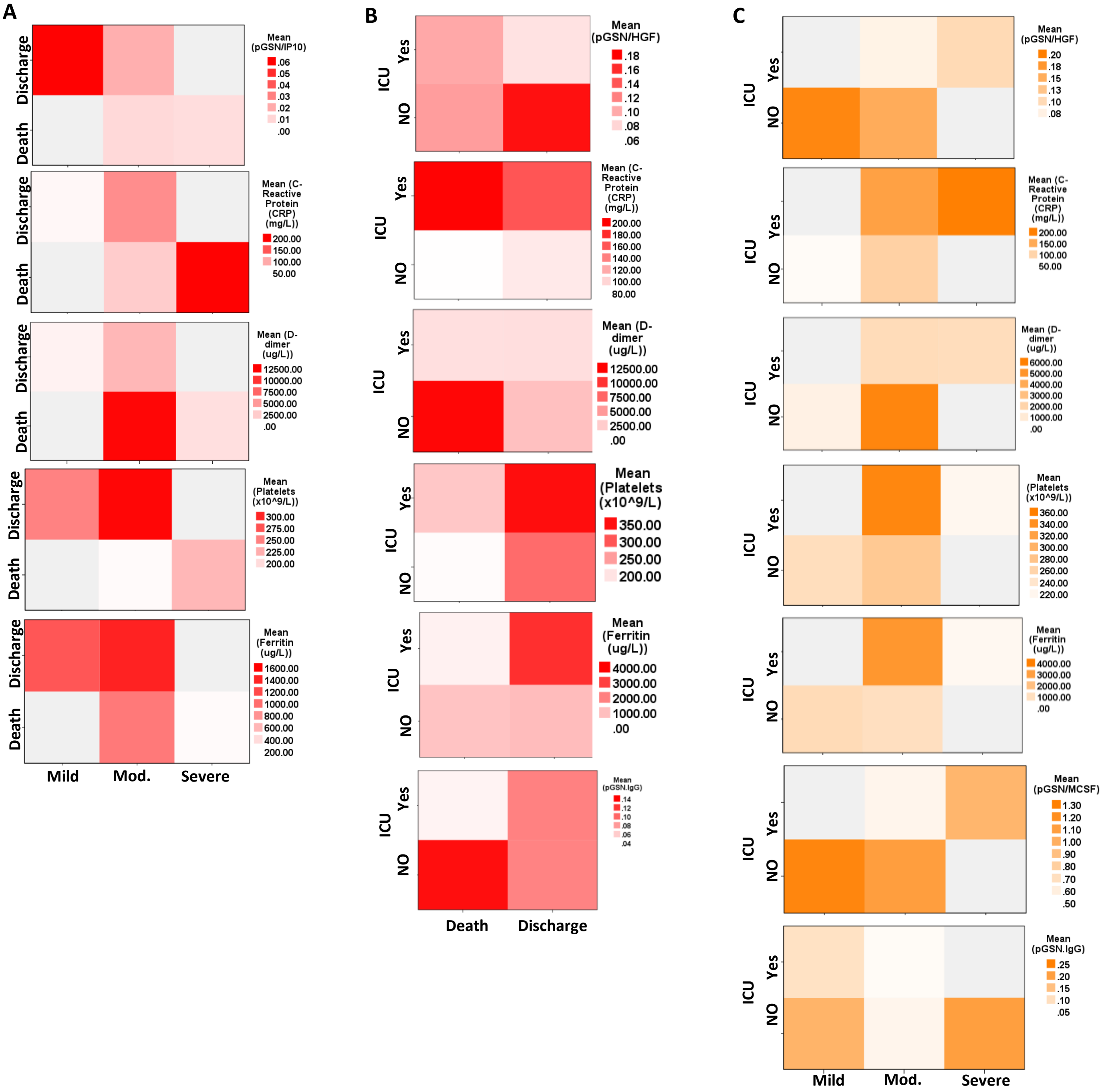


**Supp. Fig. S4. Multi-analyte panels predict more than one clinical outcomes of COVID-19.** pGSN/IP-10, pGSN/HGF, pGSN/M-CSF, CRP, platelets, Ferritin and pGSN/IgG were used in a heat map analyses to predict **(A)** disease severity and hospitalization outcomes, **(B)** ICU admissions and hospitalization outcomes and **(C)** disease severity and ICU admissions.

### Table 1: Demographics and Baseline Characteristics of COVID-19 Patients

| | | COVID-19 | | Non-COVID-19 | |
|---|---|---|---|---|---|
| | | Count | Mean | Count | Mean |
| **Hospitalization outcomes at the end of follow-up** | **Deceased** | 7 | | 0 | |
| | **Discharged** | 21 | | 0 | |
| **BMI (Kg/m2)** | | | 30.51 | | . |
| **ICU Admittance** | **No** | 23 | | 0 | |
| | **Yes** | 5 | | 0 | |
| **Age (yrs)** | | | 63.93 | | 41.67 |
| **Gender** | **Female** | 6 | | 7 | |
| | **Male** | 22 | | 5 | |
| **COVID Disease Severity** | **No infection** | 0 | | 12 | |
| | **Mild** | 7 | | 0 | |
| | **Moderate** | 18 | | 0 | |
| | **Severe** | 3 | | 0 | |
| **Days from first positive COVID-19 Test** | **2** | 2 | | 0 | |
| | **3** | 7 | | 0 | |
| | **4** | 4 | | 0 | |
| | **5** | 1 | | 0 | |
| | **6** | 1 | | 0 | |
| | **7** | 4 | | 0 | |
| | **8** | 1 | | 0 | |
| | **9** | 2 | | 0 | |
| | **11** | 1 | | 0 | |
| | **13** | 1 | | 0 | |
| | **14** | 1 | | 0 | |
| | **15** | 1 | | 0 | |
| | **17** | 1 | | 0 | |
| | **20** | 1 | | 0 | |
| | **3** | 1 | | 0 | |
| **Days from symptom onset** | **4** | 2 | | 0 | |
| | **5** | 1 | | 0 | |
| | **6** | 1 | | 0 | |
| | **7** | 3 | | 0 | |
| | **8** | 2 | | 0 | |
| | **9** | 4 | | 0 | |
| | **11** | 4 | | 0 | |
| | **12** | 2 | | 0 | |
| | **13** | 2 | | 0 | |
| | **15** | 1 | | 0 | |
| | **16** | 3 | | 0 | |
| | **20** | 1 | | 0 | |
| | **22** | 1 | | 0 | |

Disease severity was defined as follows:

1. Mild (no oxygen/room air received)
2. Moderate [supplemental oxygen, not high flow or no flow listed, no intubation/mechanical ventilation – ex. No Bi-level Positive Airway Pressure (BiPap) /intubation)].
3. Severe (high flow oxygen and non-invasive or invasive mechanical ventilation – ex. Yes BiPap/intubation)
